# Initial characterisation of commercially available ELISA tests and the immune response of the clinically correlated SARS-CoV-2 biobank “SERO-BL-COVID-19” collected during the pandemic onset in Switzerland

**DOI:** 10.1101/2020.07.05.20145888

**Authors:** Hans-Michael Kaltenbach, Fabian Rudolf, Janina Linnik, Julia Deichmann, Therese Ruf, Raffaele Altamura, Edo Kapetanovic, Derek Mason, Bastian Wagner, Thomas Goetz, Lukas Mundorff, Karin Stoll-Rudin, Christina Krebs, Tanja Renz, Thomas Hochueli, Sergio Haymoz, Markus Hosch, Nadine Périat, Michèle Richert, Sergio Sesia, Daniel Paris, Carlos Beat Quinto, Nicole M. Probst-Hensch, Christoph Niederhauser, Sai Reddy, Beatrice Nickel, Miodrag Savic

**Author notes:** these authors contributed equally (Hans-Michael Kaltenbach, Fabian Rudolf).

## Abstract

**Background:** To accurately measure seroprevalance in the population, both the expected immune response as well as the assay performances have to be well characterised. Here, we describe the collection and initial characterisation of a blood and saliva biobank obtained after the initial peak of the SARS-CoV-2 pandemic in Switzerland.

**Methods:** Two laboratory ELISAs measuring IgA & IgG (Euroimmun), and IgM & IgG (Epitope Diagnostics) were used to characterise the biobank collected from 349 re- and convalescent patients from the canton of Basel-Landschaft.

**Findings:** The antibody response in terms of recognized epitopes is diverse, especially in oligosymptomatic patients, while the average strength of the antibody response of the population does correlate with the severity of the disease at each time point.

**Interpretation:** The diverse immune response presents a challenge when conducting epidemiological studies as the used assays only detect *∼*90% of the oligosymptomatic cases. This problem cannot be rectified by using more sensitive assay setting as they concomitantly reduce specificity.

**Funding:** Funding was obtained from the “Amt für Gesundheit” of the canton Basel-Landschaft, Switzerland.

## 1. Introduction

Effective host responses to viral infections, including those to coronaviruses, are driven by adaptive immunity [1]. For endemic or previously emerging coronaviruses, the antibody response has been correlated with protection from re-infection for a varying period of time. In SARS-CoV-2 infections, most studies on antibody kinetics are based on severe or hospitalized patients, even though subclinical or even oligosymptomatic patients represent the majority of cases. Less severe cases of SARS-CoV-2 (as well as other endemic CoVs) are associated with lower antibody responses, and therefore pose a challenge for accurate detection using serological assays.

However, one of the most important correlates of immunological protection is the presence of neutralizing antibodies, which is preferably measured by using functional assays with replication competent virus [2]. For COVID-19 patients, such assays are time consuming and must be conducted in Biosafety Level 3 facilities, which renders them infeasible for wide-scale testing. Some alternative functional assays are based on pseudotyped or chimeric viral particles [3], but these reagents are neither trivial to produce nor do they scale to large sample sizes. The most feasible alternative assays are therefore binding assays, such as enzyme-linked immunosorbent assays (ELISA), and preferably report on the quantity of antibodies binding to neutralizing epitopes such as the receptor binding domain (RBD) of the spike protein [4].

Current clinically approved ELISA tests either bind to the nucelocapsid (NCP) protein or (part of) the spike protein (which includes the RBD). Both of these proteins, but especially the NCP, are known to generate a strong host immune response in other beta-coronaviruses [5, 6]. In contrast, nearly all neutralising antibodies against SARS-CoV-2 discovered to date bind to the small RBD portion of the spike protein [7, 8], and currently available commercial assays indeed insufficiently predict neutralisation [2]. The presence and characterization of antibody responses in COVID-19 patients by serological assays has been described in several reports [9, 10, 11, 12, 13, 14]; a key observation is that across many patients, antibodies are detected at *∼* 10 days post-onset of symptoms.

The performance of assays is characterized by their sensitivity and specificity [15]. To-date, most commercial ELISA performance validations were obtained from biobanks relying on hospitalised patients; this positive patient cohort will likely have higher antibody levels than milder, non-hospitalized patients [16]. This is also observed for SARS-CoV-2 infections [17]. It is thus unclear if the available tests are sufficiently sensitive to also detect oligosymptomatic cases.

Here, we describe the collection and initial analysis of a blood biobank representative for the observed COVID-19 symptomatic range in the population of Switzerland. The positive cohort in this biobank consists of 341 samples obtained from participants determined to have SARS-CoV-2 (PCR-positive test) in various symptomatic and post-symptomatic stages. The negative cohorts include 115 samples obtained from PCR-negative tested participants, and 150 samples of blood donors from the 2016/17 flu season. The distribution in age and disease severity in this biobank is similar to that reported for other areas in Western Europe. For each sample, we measured the antibody response toward the NCP and S1 proteins using the ELISA tests for IgM and IgG from Epitope Diagnostics and IgA and IgG from Euroimmun, and characterized the performance of these assays. The specificity of both the Euroimmun IgG and the Epitope Diagnostics IgM assays was close to 100%, while the other two tests showed specificities of *∼* 96% and lower. The sensitivity of the IgA and IgG tests was only sufficient to detect *∼* 90% of the cases, while the IgM test only detected *∼* 50%. Previous studies reported a low and late IgM response, especially in less severe COVID-19 patients [10, 11], which might partially explain the low IgM test sensitivity. Taken together, our data indicates that the immune response in oligosymptomatic patients is diverse and ill captured with the two employed serological assays.

## 2. Materials & methods

### 2.1. Ethics statement

This study is part of the project ‘COVID-19 in Baselland Investigation and Validation of Serological Diagnostic Assays and Epidemiological Study of Sars-CoV-2 specific Antibody Responses (SERO-BL-COVID-19)’ approved by the ethics board “Ethikkommission Nordwestund Zentralschweiz (EKNZ)”, Hebelstrasse 53, 4056 Basel representative of Swissethics under the number (2020-00816).

Every participant has received a written informed consent at least 24 hours before participating in this study (attached original document in German language). The participants had to sign the written informed consent and needed to show it in order to be given access to the test facility. The participants could withdraw their participation at any time without stating any reason.

### 2.2. Collection of samples

#### 2.2.1. Blood Collection

Venous blood was taken by puncturing a disinfected cubital or similar area using a BD safety-lock system into a vacutainer. In total, 10–12 mL each for EDTA-blood serum was taken. The blood collection was performed by a medical assistant or nurse. After blood collection, the samples were either transferred to the diagnostic lab or directly processed on site in the make-shift laboratory.

#### 2.2.2. Saliva Collection

Saliva was collected non-invasively using the dedicated Salivette tubes (Sarstedt Cat. # 51.1534). In short, the participant delivered saliva into an adsorbent filter, which was then placed by the participant in the Salivette tube. After handover to the medical staff, the saliva was centrifuged on site at 4^*°*^C using 1,000*×* g for 2 min to remove cells and debris. The tube was then rapidly frozen using a salted ice-water bath and stored at *−*20^*°*^C before transporting to the lab on dry ice and storage at *−*80^*°*^C until further use.

#### 2.2.3. Plasma and PBMC isolation and cell cryopreservation

Density gradient separation was used in peripheral blood mononuclear cell (PBMC) isolation. 12 mL of fresh donor blood was received in 3*×* 4 mL plastic whole blood tubes with spray-coated K_2_EDTA BD Vacutrainer− (Becton Dickinson, Cat. # 367844). The whole blood was diluted in 1:1 ratio with 12 mL of PBS (w/o Ca^2+^ and Mg^2+^). The total volume of diluted blood (24 mL) was gently and slowly layered on 14 mL of Ficoll Lymphoprep^*T M*^ (STEMCELL, Cat. # 07861). Samples were centrifuged at 400*×* g 40 min, 22^*°*^C, no brakes. 14 mL of plasma was transferred in a 15 mL conical tube and stored at 4^*°*^C. The layer of mononuclear cells was aspirated and transferred in a 50 mL conical tube containing 25 mL of PBS (w/o Ca^2+^ and Mg^2+^). Cells were washed 300*×* g 8 min, 22^*°*^C, with brakes. Washing was repeated with an additional 25 mL of PBS (w/o Ca^2+^ and Mg^2+^). Mononuclear cells were subsequently resuspended in 1 mL of freezing media (heat inactivated FBS supplemented with 10% DMSO) and aliquoted into two 1.5 mL cryogenic tubes (Nalgene System, Thermo Scientific, Cat. # 5000-1020). The cryogenic tubes were put into freezing containers Mr.Frosty− (Thermo Scientific, Cat. # 5100-0001) and the containers were immediately placed into an *−*80^*°*^C freezer for 24 hrs, and then transferred into a liquid nitrogen tank.

#### 2.2.4. Point of care validation

To perform the point of care test (POCT) validation, a capillary blood sample was taken from each subject by puncturing the end of a finger and taking the blood with a micro pipette. Immediately after collecting, the blood was put in the lateral flow chamber of the POCT and after 15 minutes the result was visually scored as positive or negative by the medical assistant. Additionally, the tests were imaged using a Nikon D5000 camera.

#### 2.2.5. Blood donor cohort

Samples from nonremunerated blood donors originate from the Swiss cantons of Thurgau, Basel, Bern, Waadt and Geneva, and were taken during the pre-pandemic period 16th and 17th December 2016. These samples were frozen as EDTA plasmas on microtiterplates for *−*20^*°*^C.

### 2.3. ELISA analysis

The following four commercially available immunoassays were characterized in the study: the Anti-SARS-CoV-2-ELISA-IgA (Euroimmun AG, Lübeck, # EI 2606-9601 A), the Anti-SARS-CoV-2-ELISA-IgAG (Euroimmun AG, Lübeck, # EI 2606-9601 G), the EDI Novel Coronavirus COVID-19 IgM ELISA kit (Epitope Diagnostics, Inc., # KT-1033) and the EDI Novel Coronavirus COVID-19 IgG ELISA kit (Epitope Diagnostics, Inc., # KT-1032). All ELISA kits were CE and IVD labeled.

To enable a quantitative comparison between ELISA experiments, we calculated fold changes in OD relative to the assay- and run-specific cut-off values (OD_sample_/OD_cut-off_), where OD_cut-off_ = 1.1*×*OD_cal_ for both Euroimmun ELISAs, OD_cut-off_ = (1.1 + 0.18)*×*OD_NC_ for EDI IgG and OD_cut-off_ = (1.1 + 0.10)*×*OD_NC_ for EDI IgM, where OD_cal_ and OD_NC_ are calibration respectively average negative control values as described by the manufacturers. Note that fold changes are not comparable between Euroimmun and EDI ELISA test kits because the assays show large differences in dynamic range and saturation. Detailed calculation can be found in the supplementary methods.

### 2.4. Statistical analysis

Patient data and results of POCTs were originally stored in the REDCap database system of the Canton Hospital Basel-Landschaft. Results from ELISA tests were entered into Excel worksheets. All data were preprocessed and a common database created using in-house scripts in R [18]. Statistical analysis and creation of figures and tables was carried out using R; binomial confidence intervals are 95%-Clopper-Pearson intervals calculated using exactci() from package PropCIs [19]. A refresher for the calculation of specificity and sensitivity calculation can be found in the supplementary methods.

## 3. Results

### 3.1. Study design & cohorts

The goal of our study design was to collect a representative cohort of COVID-19 disease manifestation during the first wave of COVID-19 in the canton Basel-Landschaft, Switzerland. During the initial phase of the pandemic, only people in risk groups showing symptoms were tested; later, testing was extended to all people showing symptoms and 5311 people had been tested in the canton at the beginning of study recruitment, with 802 (15.1%) positive and 4509 (84.9%) negative PCR test results. The cases were mostly observed close to or in areas with a high frequency of commuting to the city of Basel, but the ratio of positive tests showed no apparent bias for or against rural communities.

All RT-PCR-tested individuals were eligible for participation except when they were *<*18 years of age, had a severely compromised immune system, were hospitalized at the time of sample collection, or were deceased. From these, 349 positive individuals committed to participating in the study, and 111 negative individuals were randomly selected. We aimed for sufficient sample size for two positive cohorts: an ‘acute’ cohort with diagnosed COVID-19 up to 12 days before study entry, and a ‘convalescent’ cohort with more than 12 days between positive diagnosis and study entry.

Individuals were continuously recruited during a 2 week window from 11. April 2020 to 22. April 2020 and visited the ‘Abklärungsstation COVID-19’ in Münchenstein, Switzerland. The medical history and the status were recorded in a doctors interview, the vital parameters were acquired, and saliva and blood samples were collected. All participants of the positive cohort were guided through the building, while the negative cohort was examined in a make-shift field hospital erected next to the building to minimize the danger of infection with COVID-19. Participant characteristics are summarized in Table 1.

**Table 1:** Characteristics of patients included in the study.

| **Cohort** | PCR pos, $\leq 7$d<br>(N = 31) | PCR pos, $> 7$d & $\leq 12$d<br>(N = 46) | PCR pos, $> 12$d<br>(N = 272) | PCR neg, $> 5$d<br>(N = 111) |
| --- | --- | --- | --- | --- |
| **Sex** |  |  |  |  |
| female | 17 (55%) | 25 (54%) | 130 (48%) | 63 (57%) |
| male | 14 (45%) | 21 (46%) | 142 (52%) | 48 (43%) |
| **Age** |  |  |  |  |
| years, median (range) | 45 (21-80) | 51 (20-80) | 51.5 (17-93) | 48 (19-87) |
| **Weight** |  |  |  |  |
| kg, median (range) | 73 (51-110) | 72 (40-109) | 76 (49-135) | 73 (47-130) |
| **Height** |  |  |  |  |
| cm, median (range) | 173.5 (157-187) | 172 (150-191) | 173 (72-198) | 172 (149-195) |
| **SpO2** |  |  |  |  |
| %, median (range) | 98 (93-99) | 98 (88-99) | 98 (85-99) | 98 (95-99) |
| **Total days ill** |  |  |  |  |
| 0–5 days | 6 (19%) | 1 (2%) | 28 (10%) | — |
| 5–10 days | 15 (48%) | 23 (50%) | 146 (54%) | — |
| 10–14 days | 10 (32%) | 22 (48%) | 98 (36%) | — |

Distribution of ages and gender for the positive cohort are similar to the age and gender structure of the canton, except for the age between 40–65 which are over-represented and *>*80 which are under-represented (Table 2 and Supplementary Figure 1). From the 349 positive participants, 35 (10%) were bedridden during the acute disease, 62 (18%) required help for their daily activities, while 244 (72%) had no restrictions. Similar distributions are reported elsewhere. All age groups were affected equally, however severe cases were more pronounced in the older population. Increasing disease severity correlated with the experienced symptoms; bedridden cases suffered approximately 10 or more days, cases requiring help *∼*10 days, and the oligosymptomatic cases between 5 to 15 days.

**Table 2:** Age structure of study cohort stratified by severity and by duration of symptoms. Last row shows distribution of ages 15+ in canton of Basel-Land in 2019 for reference.

\*Note that our study cohort does not include minors with less than 18 years of age.
|  | Age groups |  |  |  |  |  |  |
| --- | --- | --- | --- | --- | --- | --- | --- |
|  | 15–19* | 20–29 | 30–39 | 40–49 | 50–64 | 65–79 | 80+ |
| **Overall** | 5 (1.1%) | 55 (12%) | 64 (13.9%) | 99 (21.5%) | 159 (34.6%) | 68 (14.8%) | 10 (2.2%) |
| **Severity of illness** |  |  |  |  |  |  |  |
| Not ill | 1 (20%) | 13 (24%) | 23 (36%) | 24 (24%) | 29 (18%) | 18 (26%) | 3 (30%) |
| No restrictions | 4 (80%) | 36 (65%) | 34 (53%) | 53 (54%) | 91 (57%) | 28 (41%) | 4 (40%) |
| Help needed | 0 (0%) | 6 (11%) | 6 (9%) | 18 (18%) | 22 (14%) | 9 (13%) | 1 (10%) |
| Bedridden | 0 (0%) | 0 (0%) | 1 (2%) | 4 (4%) | 17 (11%) | 13 (19%) | 2 (20%) |
| **Total days ill** |  |  |  |  |  |  |  |
| 0–5 days | 0 (0%) | 7 (17%) | 5 (12%) | 7 (9%) | 11 (8%) | 5 (10%) | 0 (0%) |
| 5–10 days | 3 (75%) | 19 (45%) | 24 (59%) | 39 (52%) | 72 (55%) | 24 (48%) | 3 (43%) |
| 10–15 days | 1 (25%) | 16 (38%) | 12 (29%) | 29 (39%) | 47 (36%) | 21 (42%) | 4 (57%) |
| Baselland (2019) | 13924 (5.6%) | 30882 (12.4%) | 35666 (14.3%) | 39190 (15.7%) | 65017 (26.1%) | 45175 (18.1%) | 19054 (7.7%) |

**Figure 1:**
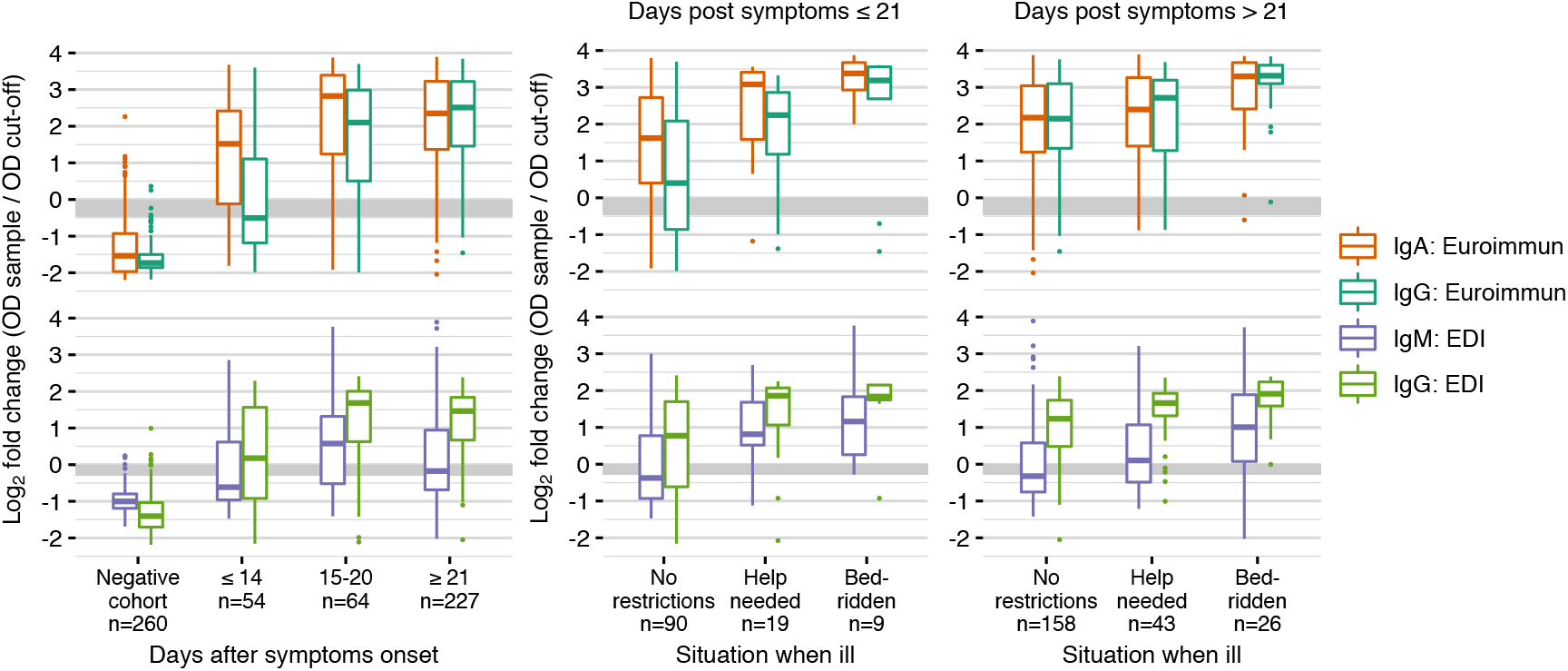
Overview on ELISA results. Epitope Diagnostics (EDI) and Euroimmun assays were performed with serum from 607 individuals. The negative cohort consists of 150 serum samples collected during the 2016/17 influenza period and 110 serum samples from PCR-negative individuals from 2020. Positive cases are stratified by days post symptoms onset (left) and additionally stratified by disease severity (right). Fold changes are defined as the ratio between measured OD and the classification cut-off OD specified by the manufacturer. Grey area indicates the range where serum samples are classified as uncertain; samples above (below) this area are classified as positive (negative).

We use two negative cohorts; (i) samples from 150 2016/17 influenza period blood donors, and (ii) our negative cohort. These cohorts should strengthen the specificity calculations, but also help to address cross-reactivity to viruses currently in circulation. However, the PCR test is prone to false negatives, and we therefore expect to find a small number of incorrectly diagnosed individuals. In line with the reported false negative rate of 15–25% [20, 21], we identified four individuals with negative PCR results but seroconverted in both Euroimmun IgG and IgA and Epitope Diagnostics (EDI) IgG. We consider these four individuals as false negative PCR results and removed them from the dataset. All results are only influenced marginally by this step.

### 3.2. Performance characteristics of ELISA tests

#### 3.2.1. Sensitivity & specificity

We performed the EDI and Euroimmun assays according to the manufacturer’s instructions on all samples. The obtained data were normalized (see Material and Methods section) to make results comparable between experiments. The reported fold-changes are defined as the ratio between measured OD and the classification cut-off OD specified by the manufacturer (Figure 1 & Supplementary Figure 2). Positive/negative classification was performed according to manufacturer’s instructions, and the assay performance was calculated therefrom.

**Figure 2:**
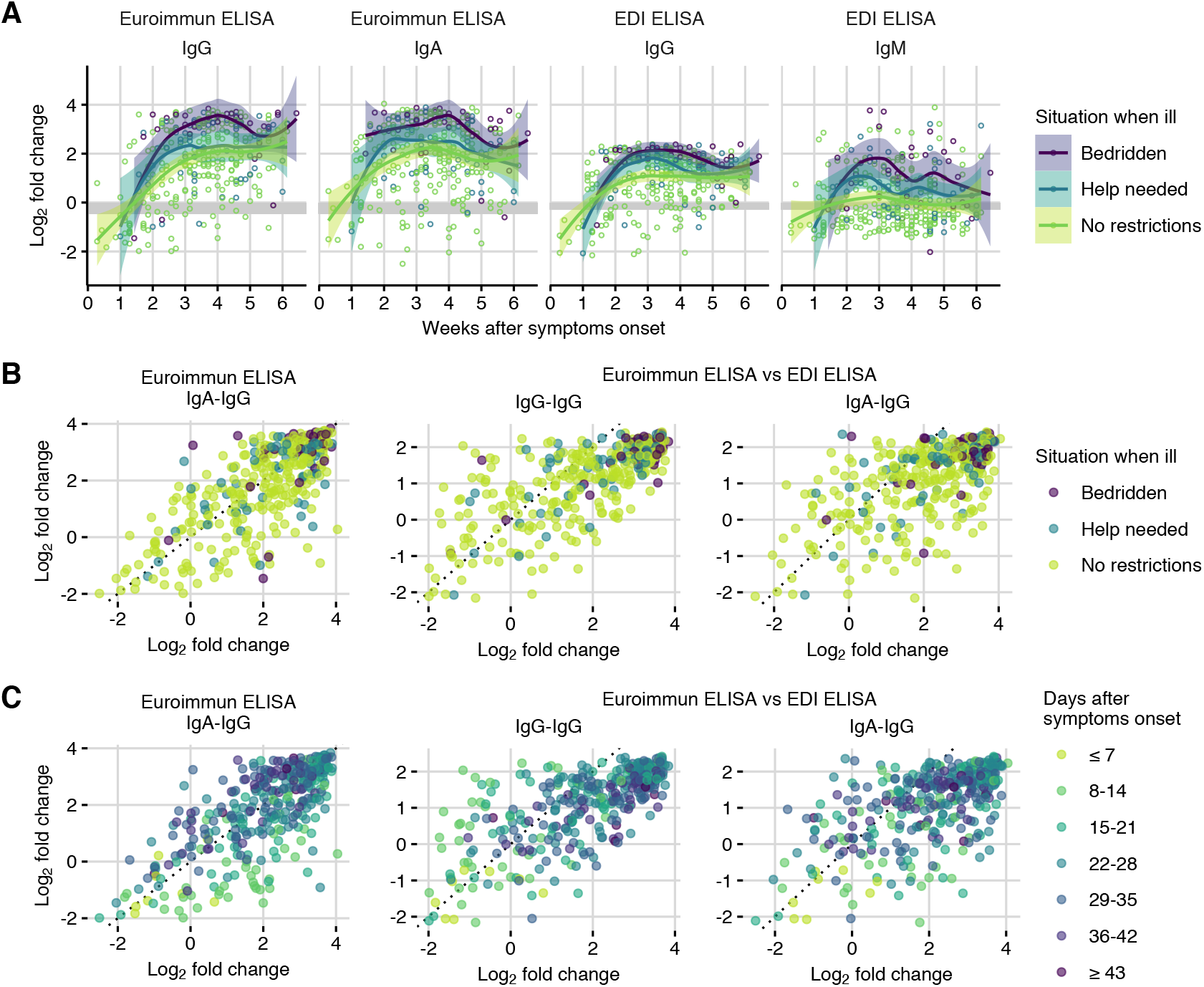
A: Log_2_-fold changes in ELISA signal for each assay by time after symptoms onset and by disease severity. Data was fitted using smoothing splines to visualize trends (figure shows fits and 95% confidence intervals). Grey area indicates the range where serum samples are classified as uncertain; samples above (below) this area are classified as positive (negative). B: Scatterplots comparing log_2_-fold changes in IgA vs IgG measured by Euroimmun ELISA (left), IgG measured by Euroimmun ELISA vs IgG measured by Epitope Diagnostics (EDI) ELISA (center), and Euroimmun IgA vs EDI IgG (right) by disease severity. C: As in B but with days after onset of symptoms. Fold-changes are defined as the ratio between measured OD and the classification cut-off OD specified by the manufacturer.

We used a patient’s assignment to the PCR-positive or PCR-negative cohort together with the corresponding ELISA test result for calculating performance characteristics for all four assays. The values considering all data are shown in the top of Table 3. We find specificities of the IgG-based assays of about 98%-99%, while IgA shows considerably lower and IgM considerably higher specificity overall. To determine potential cross-reactivity of the assays, we separately calculated the specificity from the 150 samples of the 2016 blood donor cohort (Table 3 (bottom)). Nevertheless, the histogram of all assays and data show overlap between the cohorts (Supplementary Figure 3).

**Table 3:** Top: Number true positive (TP), false positive (FP), true negative (TN), false negative (FN), sensitivity (Se) and specificity (Sp) for Epitope Diagnostics (EDI) serum and plasma samples and Euroimmun ELISA serum samples for all cohorts. All samples with uncertain result were considered negative for the analysis (number of uncertain samples shown in brackets). Column IgG 1.5*×* for Euroimmun corresponds to using a threshold of 1.5 instead of 1.1 for the OD-ratio. Bottom: specificity based on serum samples of negative 2016 blood donor cohort only.

|  | EDI |  |  |  | Euroimmun |  |  |
| --- | --- | --- | --- | --- | --- | --- | --- |
|  | Serum |  | Plasma |  | Serum |  |  |
|  | IgM | IgG | IgM | IgG | IgA | IgG | IgG 1.5× |
| TP | 161 | 287 | 176 | 280 | 304 | 291 | 272 |
| FP | 3 | 6 | 11 | 12 | 22 | 2 | 0 |
| TN | 257 (2) | 254 (7) | 100 (11) | 99 (13) | 238 (12) | 258 (2) | 260 (0) |
| FN | 184 (35) | 58 (16) | 161 (36) | 57 (14) | 41 (15) | 54 (12) | 73 (0) |
| Se [CI], % | 46.7 [41.3, 52.1] | 83.2 [78.8, 87] | 52.2 [46.7, 57.7] | 83.1 [78.6, 86.9] | 88.1 [84.2, 91.3] | 84.3 [80.1, 88] | 78.8 [74.1, 83] |
| Sp [CI], % | 98.8 [96.7, 99.8] | 97.7 [95, 99.1] | 90.1 [83, 94.9] | 89.2 [81.9, 94.3] | 91.5 [87.5, 94.6] | 99.2 [97.2, 99.9] | 100 [98.6, 100] |
| 2016 blood donors |  |  |  |  |  |  |  |
| TN | 150 (0) | 147 (3) | — | — | 136 (7) | 150 (2) | 150 (0) |
| FP | 0 | 3 | — | — | 14 | 0 | 0 |
| Sp [CI], % | 100 [97.6, 100] | 98 [94.3, 99.6] | — | — | 90.7 [84.8, 94.8] | 100 [97.6, 100] | 100 [97.6, 100] |

The IgG- and IgA-based assays show overall sensitivities above 87%, while the sensitivity of IgM is extremely low at slightly above 50%. However, the different types of antibodies act at different stages of the immune response. We therefore stratified the calculation of assay sensitivities by days after onset of symptoms into three categories: 14 days or less, 15 to 20 days, and 21 days or more (Table 4 & Supplementary Figure 4). Sensitivities increase with days after onset of symptoms for both IgG assays from about 50% to 95%. The sensitivity of the IgM assay remains low for all strata, while IgA already shows substantially higher sensitivities than all other assays for less than 14 days after onset of symptoms. Specifically, sensitivity is low for the EDI Diagnostics IgM assay, reaching a maximum of 64% for two to three weeks after onset of symptoms. Both IgG assays show overall sensitivities below 90%, but sensitivities increase to 95% or higher three weeks after onset of symptoms. The Euroimmun IgA sensitivity is highest among the four assays for less than two weeks after onset of symptoms with 82% and also reaches about 95% after three weeks or more after onset of symptoms.

**Table 4:** Sensitivity of EDI (top) and Euroimmun (bottom) ELISA stratified by days after onset of symptoms. All samples with uncertain result were considered negative for the analysis (number of uncertain samples shown in brackets).

| EDI ELISA |  |  |  |  |  |  |
| --- | --- | --- | --- | --- | --- | --- |
|  | IgM |  |  | IgG |  |  |
| | $\leq 14$ days | 15–20 days | $\geq 21$ days | $\leq 14$ days | 15–20 days | $\geq 21$ days |
| TP | 20 | 32 | 109 | 29 | 44 | 214 |
| FN | 34 (5) | 20 (4) | 130 (26) | 25 (2) | 8 (1) | 25 (13) |
| Se [CI], % | 37 [24.3, 51.3] | 61.5 [47, 74.7] | 45.6 [39.2, 52.2] | 53.7 [39.6, 67.4] | 84.6 [71.9, 93.1] | 89.5 [84.9, 93.1] |

| Euroimmun ELISA |  |  |  |  |  |  |
| --- | --- | --- | --- | --- | --- | --- |
|  | IgA |  |  | IgG |  |  |
| | $\leq 14$ days | 15–20 days | $\geq 21$ days | $\leq 14$ days | 15–20 days | $\geq 21$ days |
| TP | 40 | 46 | 218 | 23 | 42 | 226 |
| FN | 14 (5) | 6 (1) | 21 (9) | 31 (3) | 10 (4) | 13 (5) |
| Se [CI], % | 74.1 [60.3, 85] | 88.5 [76.6, 95.6] | 91.2 [86.9, 94.5] | 42.6 [29.2, 56.8] | 80.8 [67.5, 90.4] | 94.6 [90.9, 97.1] |

We performed the same analysis stratified by three levels of disease severity—‘no restriction’, ‘help needed’, and ‘bedridden’—and combined these levels with two levels for days after onset of symptoms: short (*≤* 21 days) or long (*>* 21 days). The resulting sensitivities and specificities and their 95% confidence intervals are given in Table 5. As expected, sensitivities also increase with severity of symptoms for both less and more than three weeks after onset of symptoms, but sample sizes are comparatively small for shorter time and higher severity. Notably, the IgG response is detectable in all samples of the ‘bedridden’ cohort *>* 21 days, and is then comparable to the manufacturer’s characterisation on this subset.

**Table 5:** Sensitivities stratified by severity of illness. All samples with uncertain result were considered negative for the analysis (number of uncertain samples shown in brackets).

| EDI IgM |  |  |  |  |  |  |
| --- | --- | --- | --- | --- | --- | --- |
|  | Bedridden |  | Help needed |  | No restrictions |  |
| | $\leq 21d$ | $> 21d$ | $\leq 21d$ | $> 21d$ | $\leq 21d$ | $> 21d$ |
| TP | 7 | 20 | 15 | 23 | 36 | 60 |
| FN | 2 (2) | 6 (3) | 4 (0) | 20 (6) | 54 (8) | 98 (16) |
| Se [CI], % | 77.8 [40, 97.2] | 76.9 [56.4, 91] | 78.9 [54.4, 93.9] | 53.5 [37.7, 68.8] | 40 [29.8, 50.9] | 38 [30.4, 46] |

| EDI IgG |  |  |  |  |  |  |
| --- | --- | --- | --- | --- | --- | --- |
|  | Bedridden |  | Help needed |  | No restrictions |  |
| | $\leq 21d$ | $> 21d$ | $\leq 21d$ | $> 21d$ | $\leq 21d$ | $> 21d$ |
| TP | 8 | 25 | 17 | 39 | 58 | 140 |
| FN | 1 (0) | 1 (1) | 2 (0) | 4 (2) | 32 (4) | 18 (9) |
| Se [CI], % | 88.9 [51.8, 99.7] | 96.2 [80.4, 99.9] | 89.5 [66.9, 98.7] | 90.7 [77.9, 97.4] | 64.4 [53.7, 74.3] | 88.6 [82.6, 93.1] |

| Euroimmun IgA |  |  |  |  |  |  |
| --- | --- | --- | --- | --- | --- | --- |
|  | Bedridden |  | Help needed |  | No restrictions |  |
| | $\leq 21d$ | $> 21d$ | $\leq 21d$ | $> 21d$ | $\leq 21d$ | $> 21d$ |
| TP | 9 | 25 | 18 | 40 | 71 | 141 |
| FN | 0 (0) | 1 (0) | 1 (0) | 3 (2) | 19 (6) | 17 (7) |
| Se [CI], % | 100 [66.4, 100] | 96.2 [80.4, 99.9] | 94.7 [74, 99.9] | 93 [80.9, 98.5] | 78.9 [69, 86.8] | 89.2 [83.3, 93.6] |

| Euroimmun IgG |  |  |  |  |  |  |
| --- | --- | --- | --- | --- | --- | --- |
|  | Bedridden |  | Help needed |  | No restrictions |  |
| | $\leq 21d$ | $> 21d$ | $\leq 21d$ | $> 21d$ | $\leq 21d$ | $> 21d$ |
| TP | 7 | 25 | 16 | 42 | 53 | 148 |
| FN | 2 (0) | 1 (1) | 3 (0) | 1 (0) | 37 (7) | 10 (4) |
| Se [CI], % | 77.8 [40, 97.2] | 96.2 [80.4, 99.9] | 84.2 [60.4, 96.6] | 97.7 [87.7, 99.9] | 58.9 [48, 69.2] | 93.7 [88.7, 96.9] |

#### 3.2.2. Reproducibility and linearity

Intra-assay variability was determined by calculating the mean, standard deviation, and coefficient of variation of the measured OD ratios based in 5 replicate measurements of 8 samples with values ranging from at or below the threshold limit to about the 75% quartile in the two IgG and IgA assays (Supplementary Table 5 & Supplementary Figure 6). In general, reproducibility was high with coefficients of variation of 5% or smaller in samples well above the threshold. This variation increased to *∼* 10% at or below the threshold. It is trivial but important to state that, at the threshold, this variability can lead to different classification in each repetition. Last, different samples show different variabilities in the two IgG assays, possibly indicating the difference between the binding epitope and assay manufacturing.

We investigated the assay linearity using two-fold serially diluted serum samples from 5 individuals with high OD values for IgG (Supplementary Figure 11). Both Euroimmun assays showed a good linearity over a dilution range of 64-fold (2^6^ dilutions), while the IgG Epitope Diagnostics (EDI) assay is linear only in a 16-fold (2^4^) dilution range. Additionally, both IgG assays show good agreement along the dilutions (Supplementary Figure 10), suggesting that—after normalization and scaling—the IgG assays give comparable quantitative results. No data of sufficient quality was acquired for a discussion of the IgM assay (Supplementary Figure 11). Additionally, we found that the IgG, but not the IgM, assay from EDI is sensitive to haemolysis (Supplementary Figure 12).

### 3.3. Kinetics of seroconversion

Our positive cohort can give insight into the kinetics of seroconversion at the population level for different antibody types and epitopes and a broad range of disease manifestations. The two ELISA tests employ different epitopes: the EDI test recognizes the SARS-CoV-2 nucleocapsidprotein and uses IgG and IgA while the Euroimmun test recognizes the S1 portion of the spike protein using IgG and IgM.

For all antibody types, the strength of the response over time correlated with severity of the symptoms (Figure 2A). We only measured a well discernible IgM response in the “bedridden” group and a slight response in the “help needed” group, while it was largely absent from the “no restriction” group. On the other hand, the IgA and IgG response is measurable in most samples. The average response is higher when “bedridden”, while similar antibody response levels are observed for “no restriction” and “help needed”.

In most viral host responses, the earliest measurable response is IgM followed by IgG and IgA. Surprisingly, but in line with other reports for SARS-CoV-2 [10, 11] as well as observations for SARS and MERS, all three antibody types reacted within a similar timeframe. The earliest detection, within a week of symptom onset, was for IgA, followed by IgM and IgG both detected from week 2. IgM then peaked 2–3 weeks and IgA 3–4 weeks post symptom onset, while no decline for IgG was measured. The temporal order of these peaks can be expected based on the half-life time of the different antibody types in the blood.

These observations are further substantiated from observing the responses on the level of the individuals. Most interestingly, we find a robust IgG response against the NCP early on, whereas the response to the spike protein seems to be more prevalent at later time points (Figure 2A). However, neither IgG response is significantly different depending on the severity of the disease. The IgA response against the spike protein is produced rapidly in the absence of an IgG response but subsides at later time points when the IgG response becomes dominant.

## 4. Discussion

Here, we describe the collection and initial characterisation of a blood bank from the affected population of the canton Basel-Landschaft, Switzerland. Almost 50% of the people fallen ill with COVID-19 participated in this study and the biobank is therefore exemplary for an outbreak in a western European community. In particular, the COVID-19 severity and symptoms in the biobank samples closely follow the reported distributions elsewhere. We might have slightly biased our cohorts toward less severe cases, by excluding samples from actively hospitalized or deceased patients. Since only 11 cases remained in the hospital at the time of recruitment, this bias is likely small. Overall, we consider the largely representative sample a strength of our biobank, as it allows us to estimate the kinetics of seroconversion and assay performance characteristics on samples closely resembling those expected in population-wide studies for sero-prevalence.

We find that both specificity and sensitivity are below the manufacturer’s specifications for all four tests, and observe similar sensitivities only for our ‘bedridden’ group. This indicates that characterizations by the manufacturers are likely based on ‘severe’ cohorts, which limits their applicability and impairs planning of sero-prevalence studies. We also find that the IgA and IgG assays allow for quantification within a relatively narrow range: the linear range of the Euroimmun assay covers approximately a 64-fold dilution range and the EDI assay a 16-fold dilution range. When calculating sensitivities and specificities with our cohort, the IgA test shows the highest sensitivity, followed by the two IgG assays. The poor sensitivity of the IgM test is either due to the absence of a dedicated IgM response [22] in line with previous reports or poor test performance. The highest specificity is achieved with the Euroimmun IgG test. The specificity could be further boosted by a higher cut-off as previously reported, however accompanied by a concurrent drop in sensitivity (Supplementary Tables 14, 15, 16).

The average strength of the antibody response, as measured by the ratios, correlates with the severity of the disease at each time point. However, the differences vanish the longer post-symptoms onset the individuals are, and the measured amount in an individual is not predictive for the severity of the disease. Surprisingly, but similar to earlier reports, we measured that the temporal order of the response is shifted. IgA is measured within 1–2 weeks after symptoms onset, followed by the IgG response at week 2–3, while a clear IgM response is absent. Especially at earlier time points, the IgA response is stronger than the IgG response as measured using the S1 spike protein epitope in the Euroimmun assay. To fully understand what the obtained values reflect and how they correlate with diseases and protection from disease, further analyses are planned including measuring the strength to neutralize pseudo-typed viral particles and analysing the individual antibodies by sequencing the PBMCs. However, our current data help to understand the limitations of the current antibody tests on the market and serve as the basis to interpret ongoing studies on other lab assay characterization, POCT performances and PBMC sequencing.

## Data Availability

Data are available upon request

## Acknowledgement

This study was sponsored by Jürg Sommer, head of the “Amt für Gesundheit”, and the logistics of the sample collection were provided by the crisis staff and the civil protection service of the canton Basel-Landschaft. FR is funded by the NCCR ‘Molecular Systems Engineering’. Funding for JD from the two Cantons of Basel through project grant PMB-01-17 granted by the ETH Zurich is acknowledged.

Funding for TR, DP, BN Funding for MS

